# Analysis of Frailty in Peritoneal Dialysis Patients Based on Logistic Regression Model and XGBoost Model

**DOI:** 10.1101/2024.07.29.24311190

**Authors:** Qi Liu, Guanchao Tong, Qiong Ye

**Affiliations:** Wenzhou Central Hospital, 252 Baili East Rd, Lucheng, Wenzhou, Zhejiang Province 325000, China; Wenzhou-Kean University, 88 Daxue Rd, Ouhai, Wenzhou, Zhejiang Province 325060, China; Kean University, 1000 Morris Avenue, Union, New Jersey 07083, USA

**Keywords:** peritoneal dialysis, frailty, FRAIL scale, Logistic Regression, XGBoost model

## Abstract

**Purpose:** The aim of this study was to establish a model that would enable healthcare providers to use routine follow-up measures of peritoneal dialysis to predict frailty in those patients.

**Design:** A cross-sectional design with Logistic regression and XGBoost machine learning algorithms analysis.

**Methods:** One hundred and twenty-three cases of peritoneal dialysis patients who underwent regular follow-up at our center were included in this study. We use the FRAIL scale to confirm the frailty of the patients. Clinical and Laboratory data were obtained from the peritoneal dialysis registration system. Factors associated with patient Frailty were identified through regularized logistic regression and validated using an XGBoost model. The final selected variables were in-cluded in the unregularized Logistic Regression to construct the model

**Findings:** A total of 123 patients were reviewed in this study, with an average age of 61.58 years, and the median dialysis Duration was 38.5(18.07,60.53) months. 39 patients (31.71%) were female, 54 PD patients (43.9%) were classified as frail. Age, Ferritin, and TCH are the top three im-portant features labeled by the XGBoost. The results are consistent with the regularized logistic regression.

**Conclusions:** In this study, age, total cholesterol, and ferritin are the most important features associated with the frailty in peritoneal dialysis patients. This model can be used to predict frailty status and help health monitoring of peritoneal dialysis patients.

**Clinical Evidence:** Logistic regression and XGBoost machine learning algorithms can be used to construct a predictive model of frailty in peritoneal dialysis patients. The model could provide doctors with an objective tool to find frailty in peritoneal dialysis patients. As the data is obtained from routine examinations, the prediction model will not bring additional burden to the work of doctors or nurses.

## INTRODUCTION

Peritoneal dialysis can bring many benefits to patients with end-stage renal disease and prolongs their lives. However, it also has disadvantages, such as infections, metabolic and mechanical complications, risk of inadequate dialysis or limited possibilities to increase adequacy, malnutrition, long-term viability, psychological problems related to indwelling abdominal dialysis catheters, and fatigue from continuous use, particularly among elderly people[1].

Frailty is a commonly used term to denote a multidimensional syndrome of loss of reserves (energy, physical ability, cognition, health), which gives rise to vulnerability[2]. Frailty is highly prevalent in older patients with dialysis-dependent kidney failure[3].

The causes of frailty in PD patients are a little more complex. The buildup of toxins in uremia can cause loss of appetite and fatigue. Among those undergoing dialysis, particularly peritoneal dialysis (PD), challenges such as abdominal distention and bloating can exacerbate the loss of appetite. This diminished appetite, combined with dietary restrictions and reduced physical activity, leads to protein-energy wasting, muscle breakdown, and the onset of sarcopenia[6], [7]. Elevated levels of uremic toxins in circulation and disruptions in acid-base balance contribute to chronic inflammation and hinder the effectiveness of anabolic hormones, ultimately resulting in catabolism and impaired energy utilization[6]. Moreover, dialysis removes uremic toxins and restores homeostasis, thus theoretically improving weakness[7]. The interaction between malnutrition, inflammation, and frailty often coexist and have a combined effect on patient outcomes[8].

In patients with chronic kidney disease, both before or after dialysis initiation, frailty has been linked with increased risks of falls, hospitalization, cognitive dysfunction, and mortality[7], [9]. Early identification may enable timely intervention to prevent adverse health outcomes in this group of patients.

A significant amount of research has employed statistical learning and machine learning algorithms to address medical problems and identify relationships between various factors through patient observation and indicators of different diseases[10]. Machine learning can process large data sets and then efficiently process this data into clinical knowledge that enables physicians to prepare and deliver treatments, ultimately improving outcomes[11]. Based on clinical data, machine learning models can diagnose aphasia, and urinary tract infections, and predict breast cancer, among others, providing clinicians with a “second opinion”.

This study aims to analyze the relationship between frailty and various indicators (especially nutritional related) through the observation of patients with regular peritoneal dialysis follow-up in our center by using logistic regression and extreme Gradient Boost (XGBoost) algorithms and try to find out the important factors of frailty in patients with peritoneal dialysis, to intervene in the early stage and improve the quality of life of patients.

## MATERIALS AND METHODS

### Participants

A total of 123 patients with peritoneal dialysis followed up for more than 1 year were included in the study. The inclusion criteria were patients on maintenance PD and age >18 years old, with basic listening, speaking, reading, and understanding skills. The exclusion criteria were patients with critical illness, unable to cooperate with the required actions, unable to understand the content of the scale, patients with acute infection, tumor patients, etc.

This study was approved by the Wenzhou Central Hospital Human Research Ethics Committee, and prior written consent was obtained from the Participants.

### Data collection

The study started with registration on September 02, 2023. Trained and qualified nurses explained the study’s purpose to patients and obtained their informed consent. They administered the scale questions by reading them and recording the patient’s responses. Researchers then reviewed the completed scales to ensure data accuracy and consistency.

Clinical and Laboratory data were obtained from the peritoneal dialysis registration system.

### Assessment of Frailty

The FRAIL scale includes 5 components: Fatigue, Resistance, Ambulation (slow walking speed), Illness, and Loss of Weight (5% or more in the previous year). Frail scale scores range from 0–5 (i.e., 1 point for each component; 0=best to 5=worst) and represent frail (3–5), pre-frail (1–2), and robust (0) health status[12]. The FRAIL scale has been recommended by the latest international guidelines as the best screening tool for Frailty[13]. In a study of HD (hemodialysis) patients, the FRAIL scale proved to be an easy-to-apply tool, identifying a high prevalence of frailty and being a predictor of hospital admission[14]. In this study, we combine frail and pre-frail, classify 1-5 as frail, and 0 as non-frail status.

### Logistic Regression model

Logistic regression is a widely used classification algorithm that uses a linear model to predict the probability of a binary outcome[24]. The model of the binary logistic regression is a linear regression of the log odds which can be expressed as[15]:

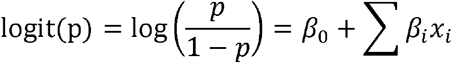

It is a simple and effective way to model binary data, but it can sometimes suffer from overfitting. The loss function of the Overfitted models will fail to replicate in future samples, thus creating considerable uncertainty about the scientific merit of the finding[16]. Regularization is a technique used to avoid overfitting in machine learning models. The lasso, ridge, and elastic net are popular regularized regression models for supervised learning[17]. Elastic Net regularization combines L1 (Lasso) and L2 (Ridge) regularization by adding a penalty term to the objective function that is a combination of the absolute value and square of the coefficients. The loss function of the elastic net model can be expressed as[18]:

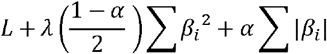

In the loss function, the value of the α controls the mix of the lasso and ridge regression. When α = 0, it becomes the pure ridge regression, and when α = 1, it is the pure lasso regression.

### XGBoost Model

XGBoost was mainly designed for speed and performance using gradient-boosted decision trees. It represents a way for machine boosting, or in other words, applying boosting to machines[19]. It is based on the sparsity-aware algorithm and weighted quantile sketch, which can converge the weak learners step-wisely into the ensemble to form a strong learner. Overfitting also can be reduced largely in the XGBoost model through parallel calculation and regularization lifting technology[20].

### Data Analysis

IBM SPSS Statistics 25 and R-Studio software (version 4.3.2) were used for data analysis. The Continuous data were expressed in different ways according to the results of normal tests, mean ± standard deviation, or median (interquartile range). The count data were expressed as the percentage of cases. Regularized Logistic Regression analysis was used to filter the effective variables for the frailty. The variables with non-zero regression coefficients would be included in the final model. The importance of the prediction model variables was ranked by XGBoost, by which the results of Regularized Logistic Regression analysis would be verified. The final selected variables were included in the unregularized Logistic Regression to construct the model (Figure 1).

**Figure 1.**
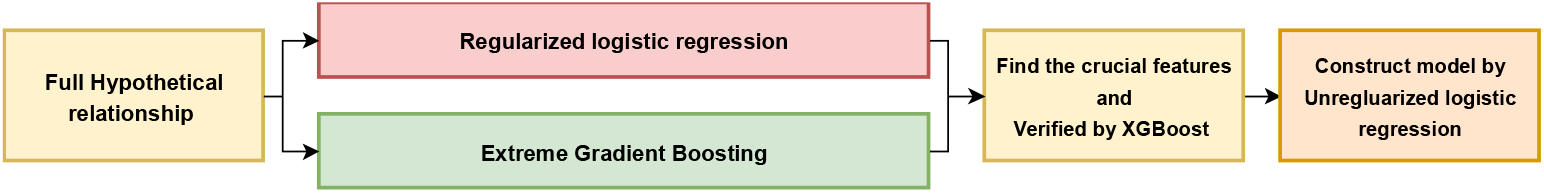
Flow Chart of the procedure: Regularized logistic regression analysis and the XGBoost model were used to screen variables. The three features significantly related to the frail were filtered by regularized logistic regression and are the top three important variables in the XGBoost importance. The results from regularized logistic regression and XGBoost are consistent. The unregularized logistic regression was used to construct the prediction model for the frail.

## RESULTS

### Participants and Baseline Frailty Status

A total of 123 patients were reviewed in this study, with an average age of (61.58± 12.8) years, and the median dialysis Duration was 38.5(18.07,60.53) months. 39 patients (31.71%) were female, 54 patients with original kidney disease (43.9%) were hypertensive nephropathy, and 54 PD patients (43.9%) were classified as frail(Table 1).

**Table 1.** Patient demographics. There are a total of one hundred and twenty-three patients in the dataset, of which forty-five Peritoneal Dialysis (PD) patients were frail.

| Variable | All patients | Not frail | Frail |
| --- | --- | --- | --- |
| Number | 123 | 69 | 54 |
| Gender (%) |  |  |  |
| Male | 84(68.29) | 50(72.46) | 34(62.97) |
| Female | 39(31.71) | 19(27.54) | 20(37.04) |
| Education level (%) |  |  |  |
| Never go to school | 19(15.45) | 9(13.04) | 10(18.52) |
| Primary school | 54(43.90) | 27(39.13) | 27(50.00) |
| Junior school | 33(26.83) | 20(28.99) | 13(24.07) |
| High school | 13(10.57) | 11(15.94) | 2(3.70) |
| College degree or above | 4(3.25) | 2(2.90) | 2(3.70) |
| Primary Diagnosis of Renal Disease (%) |  |  |  |
| Hypertension | 54(43.90) | 34(49.28) | 20(37.04) |
| Diabetes | 50(40.65) | 22(31.88) | 28(51.85) |
| Chronic Glomerulonephritis | 15(12.20) | 9(13.04) | 6(11.11) |
| Others | 4(3.25) | 4(5.80) | 0(0) |
| Age(year) | 61.58±12.56 | 57.94±11.78 | 66.56±11.68 |
| Dialysis Duration (months) | 38.50(42.46) | 36.63(43.74) | 38.00(36.72) |
| Vitamin D(nmol/L) | 42.20(20.90) | 41.50(21.60) | 42.55(20.50) |
| HB(Hemoglobin)(g/L) | 113.20±17.32 | 114.00±19.22 | 112.44±14.79 |
| Albumin(g/L) | 34.7(4.8) | 34.75(4.8) | 34.1(5.0) |
| TCH(Total Cholesterol)(mmol/L) | 4.26(1.54) | 4.38(1.96) | 4.04(1.62) |
| TG(Triglyceride)(mmol/L) | 2.09(1.59) | 2.05(1.82) | 2.08(1.54) |
| Ca(mmol/L) | 2.32(0.21) | 2.30±0.20 | 2.33±0.17 |
| PTH(Parathyroid Hormone)(pg/mL) | 198.50(330.75) | 200.00(300) | 193.50(362) |
| Prealbumin(mg/L) | 335.00±143 | 350.85±102.80 | 345.98±108.66 |
| Iron saturation | 0.23(0.17) | 0.21(0.19) | 0.25(0.16) |
| Ferritin(ug/L) | 78.70(119.40) | 99.80(163.7) | 63.35(97.2) |
| β2-microglobulin(mg/L) | 36.55(20.25) | 38.64(18.05) | 34.42(23.99) |
| Transferrin(g/L) | 2.49(0.93) | 2.45(1.07) | 2.54(0.79) |

### Regularized Logistic Regression Results and Verified by XGBoost

Using the Regularized Logistic Regression algorithm with optimal parameters (Alpha=0.5, lambda=0.15), three significant factors associated with frailty occurrence in PD patients, each with non-zero coefficients, were selected from clinical data. As shown in the Table 2, the three most important factors are Age, Total Cholesterol, and Ferritin.

**Table 2.** The Importance and the Coefficients for the Regularized Model. We find three variables are significantly related to the frailty occurrence in PD patients: Age, Total Cholesterol, and Ferritin.

| Variable | Variable Importance | Coefficients* |
| --- | --- | --- |
| Gender | 0 | 0 |
| Age | 0.0252664469 | 0.0252608776 |
| Dialysis Duration | 0 | 0 |
| Education.Level | 0 | 0 |
| Protopathy | 0 | 0 |
| Vitamin D | 0 | 0 |
| HB | 0 | 0 |
| Albumin | 0 | 0 |
| TCH | 0.0016228674 | -0.0015955269 |
| TG | 0 | 0 |
| Ca | 0 | 0 |
| PTH | 0 | 0 |
| Prealbumin | 0 | 0 |
| Iron saturation | 0 | 0 |
| Ferritin | 0.0001208403 | -0.0001204672 |
| β2-microglobulin | 0 | 0 |
| Transferrin | 0 | 0 |

\*The Intercept is -1.7843008324

Using the XGBoost algorithm with optimal parameters (max_depth = 4, verbose = 0, nrounds = 20). The factors are ranked by importance from XGBoost as shown in Table 3 and Figure 2. Table 3 shows the three measures: Gain, Cover, and Frequency in the variable importance matrix [19]. The Gain implies the relative contribution of the corresponding feature to the model. The Cover shows the relative number of observations related to the corresponding feature. Moreover, The Frequency represents the percentage of times a particular feature occurs in the model. The Gain is the most relevant attribute for interpreting the relative importance of each feature[19]. Figure 2 shows the visualization of the Gain of the different features.

**Table 3.** The Importance Matrix of the Variables from XGBoost There are three measures: Gain, Cover and Frequency in the variable importance matrix The Gain is the most relevant attribute for interpreting the relative importance of each feature.

| Variable | Gain | Cover | Frequency |
| --- | --- | --- | --- |
| Age | 0.17481068 | 0.08966518 | 0.09302326 |
| Ferritin | 0.15382618 | 0.06178086 | 0.07441860 |
| TCH | 0.08037195 | 0.04529906 | 0.06511628 |
| HB | 0.07744565 | 0.06675650 | 0.06976744 |
| Vitamin D | 0.07142420 | 0.07784804 | 0.06511628 |
| TG | 0.06544205 | 0.04840883 | 0.07441860 |
| Iron.saturation | 0.05993071 | 0.06914067 | 0.06046512 |
| PTH | 0.05386862 | 0.04312221 | 0.04186047 |
| Ca | 0.05379215 | 0.11827511 | 0.07906977 |
| β2-microglobulin | 0.05291816 | 0.09049445 | 0.06511628 |
| transferrin | 0.03860538 | 0.03130507 | 0.05581395 |
| Albumin | 0.03137946 | 0.05763450 | 0.05116279 |
| Prealbumin | 0.03117914 | 0.06748212 | 0.06046512 |
| Protopathy | 0.02292476 | 0.06530528 | 0.02790698 |
| Dialysis Duration | 0.02013171 | 0.02726236 | 0.05116279 |
| Education Level | 0.00622742 | 0.01585985 | 0.02790698 |
| Gender | 0.00572177 | 0.02435990 | 0.03720930 |

**Figure 2.**
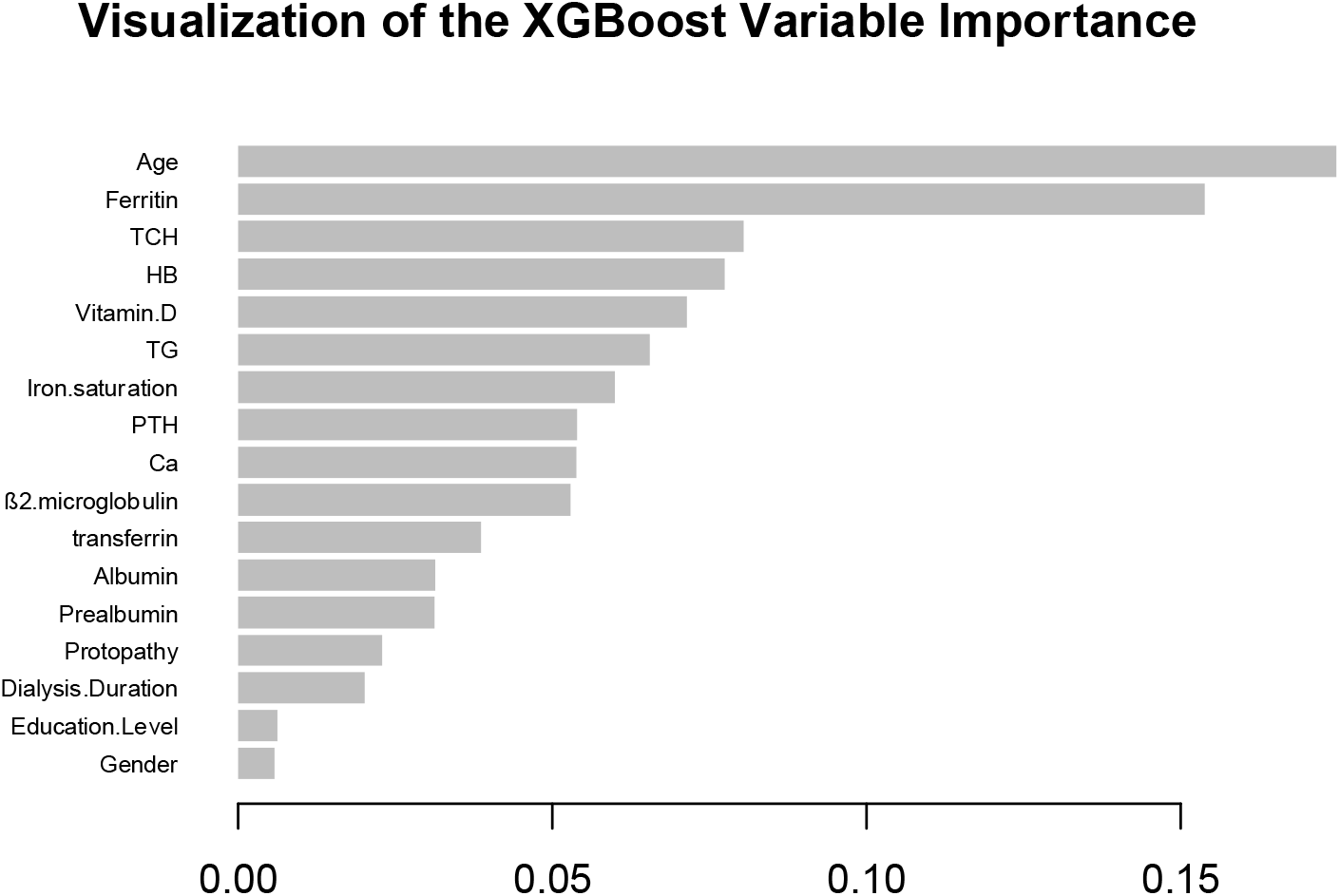
XGBoost Variable Importance Plot: The Variable importance (Gain) of the XGBoost. Age, Ferritin, and TCH are the top three important features labeled by the XGBoost. The results are consistent with the regularized logistic regression.

The top three features in the Gain column, Age, Total Cholesterol, and Ferritin, are consistent with the results of the regularized logistic regression model.

### Unregularized Logistic Regression

After applying the regularized logistic regression and being verified by XGBoost, we determined that age, total cholesterol, and ferritin are the three significant factors associated with the occurrence of frailty in PD patients. Then, we use the unregularized Logistic Regression to obtain the relationship between the three variables and the occurrence of frailty.

Based on the results of Unregularized Logistic Regression, a frailty prediction model for PD patients was established using three independent predictive factors: age, total cholesterol (TCH), and ferritin. The prediction model is as follows:

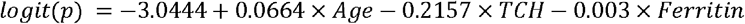

, where logit(p) is equal to the log-odds of the p, which is 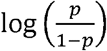.

In the model above, the p is the probability of frailty. The model above shows that increasing one unit (year) of the age will result in a 0.0664 increase in logit(p), which means there is a 6.8% increase in the odd of probability of frailty. Increasing one unit (mmol/L) of the TCH will result in a 0.2157 decrease in logit(p), which means there is a 19.4% decrease in the odd of the probability of frailty. Increasing one unit (ug/L) of the Ferritin will result in a 0.003 decrease in logit(p), which means there is a 0.30% decrease in the odd of the probability of frailty.

The area under the receiver operating characteristic curve (AUC) for the unregularized logistic regression model is 0.736 as shown in Figure 3. The model shows a positive association between age and frailty, while total cholesterol and ferritin are nagetively associated with the development of frailty.

**Figure 3.**
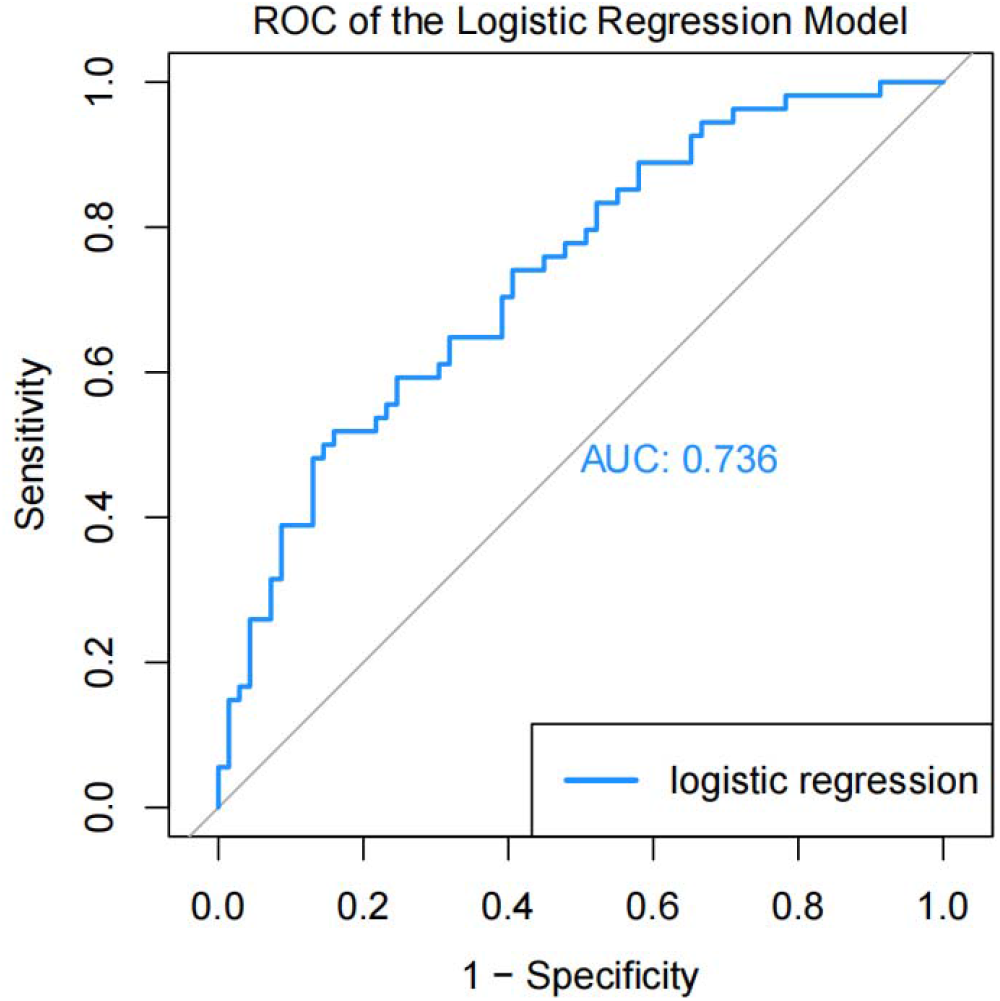
The ROC of the Unregularized Logistic Regression Model: The AUC for the unregularized logistic regression model is 0.736

## DISCUSSION

We found that 43.9% of peritoneal dialysis patients from our data had different levels of frailty based on the FRAIL scale. The main related factors for frailty are age, total cholesterol, and ferritin.

In this study, age was the most important factor for frailty in both logistic regression and importance analysis with XGBoost, which is consistent with previous studies[21], [22] (Figure 2). While the biological causative mechanisms of frailty are different from those processes causing the aging process, age is still the most important factor affecting frailty.With aging, one or more physiological systems change especially the neuro-muscular, neuroendocrine, and immune systems[23]. These changes interact and accumulate, decreasing physiological function and reserve. Frailty is thought to result when damage to multiple physiological systems exceeds a threshold so much that repair mechanisms fail to maintain system homeostasis[22], [24]. Peritoneal dialysis patients often face a lot of health challenges, such as malnutrition, metabolic disorders, and cardiovascular disease[8]. Peritoneal dialysis may affect patients’ physiological function, immune system function, mental health, and quality of life[1], [25]. The patients often have chronic diseases such as diabetes and hypertension. Peritoneal dialysis patients would be sensitive to the age-related features, such that the effect of age on frailty may be exacerbated.

Too much cholesterol could increase the risk of heart disease, stroke, and pancreatitis.However, the low cholesterol is also related to nutrition, infections, tumors, and liver disease[26], [27]. For patients on regular dialysis, the lower total cholesterol is usually due to malnutrition and inflammation[28]. A cross-sectional study in older adults found being at a high level of TCH was associated with a lower risk of frailty. A low TCH level might be the most associated with frailty risk[29]. People with lower TCH (≤5.2 mmol/l) were more likely to decline in functional status compared to others with higher TCH[27]. The complex effects of cholesterol may be related to age. A report showed high midlife TCH is a risk factor for subsequent dementia/Alzheimer’s disease, but decreasing TCH after midlife may reflect ongoing disease processes and may represent a risk marker for late-life cognitive impairment[30]. Additionally, other studies show that total TCH is inversely associated with mortality as a metabolic shift and reverse metabolism as well[31], [32]. Hypocholesterolemia was associated not only with an increased risk of mortality but also with an increased rate of complications (especially infections), clear biochemical evidence of malnutrition, and higher duration and cost of hospitalization[33]. In our study, the total cholesterol of the patients was 4.26(1.54)mmol/L, and only 15 percent were above the upper limit of normal. Because the overall TCH level is not high, this may be the reason why the lower the TCH, the higher the risk of frailty in our study.

Ferritin is an important iron storage protein in the human body and participates in the regulation of hematopoietic and immune systems. Ferritin levels can reflect the iron reserve and nutritional status of the body, and it is related to a variety of diseases[34], [35]. We found a relationship between ferritin and frailty. A cross-sectional study in a tertiary care center also showed ferritin was an independent significant predictor of frailty[36]. The effect of ferritin on frailty may be related to factors including anemia, oxygen supply, aerobic metabolism, inflammation, immune response, and malnutrition [37], [38].

As people age, frailty has long been considered a state that accelerates the risk of adverse outcomes. Frailty is more common in patients with uremia and peritoneal dialysis[7]. Identifying biomarkers associated with frailty may benefit the care of peritoneal dialysis patients. Our center’s routine follow-up of peritoneal dialysis patients includes the complete blood count (CBC), Biochemistry Panel, and other indicators. Compared to frailty assessment scales such as FRAIL and FSQ(Frailty Screening Questionnaire), which include subjective questions about fatigue, stair climbing difficulties, and community ambulation[39], [40], these biochemical indicators are easy to obtain, stable, and objectives.

Our results may help to improve the ability to identify those adults at greatest risk of frailty in the management of peritoneal dialysis patients and provide early intervention afterward. We should pay attention to the age, total cholesterol, and ferritin levels during routine examinations of peritoneal dialysis patients. Patients with frailty risk can be further examined with a frailty scale to improve the accuracy of the diagnosis (such as the FRAIL scale).

However, some indicators that have diagnostic significance for frailty in other articles were not screened out in this study (such as albumin, hemoglobin, etc.) [36], [41]. The reason may be that our sample size is too small. Therefore, in the future, we plan to expand the sample size and use big data analytics to train and refine this model, and use the model to detect the peritoneal dialysis patients. The model could provide doctors with an objective tool to diagnose frailty in peritoneal dialysis patients. As the data is obtained from routine examinations, the prediction model will not bring additional burden to the work of doctors or nurses.

## CONCLUSION

In this study, age, total cholesterol, and ferritin are the most important features associated with the frailty in peritoneal dialysis patients. This model can be used to predict frailty status and help health monitoring of peritoneal dialysis patients. In the future, we will collect more data to verify and improve the accuracy of the model.

## Data Availability

All data produced in the present study are available upon reasonable request to the authors

## Institutional Review Board Statement

The study was conducted in accordance with the Declaration of Helsinki and approved by Wenzhou Central Hospital.

## Informed Consent Statement

Written informed consent has been obtained from the patients to publish this paper.

## Data Availability Statement

Dataset available on request from the authors

